# Spatio-temporal transmissibility and dispersion of SARS-CoV-2 variants and sub-variants of concern in England

**DOI:** 10.1101/2024.08.14.24311985

**Authors:** Ben Swallow, Joshua Grier, Jasmina Panovska-Griffiths

**Affiliations:** School of Mathematics and Statistics, University of St Andrews, St Andrews, KY16 9LZ, UK; University College, University of Oxford, OX1 4BH, Oxford, UK; The Queen’s College, University of Oxford, OX1 4AW, Oxford, UK; Pandemic Sciences Institute, University of Oxford, OX3 7LF, Oxford, UK

## Abstract

The SARS-CoV-2 pandemic was characterised by continual emergence of variants. For improved future pandemic preparedness it is important to understand whether each successive variant has been more infectious and more widely spread. In this paper, we used genetic sequencing data from the COVID-19 Genomics UK Consortium in England and robust statistical models to quantify the transmissibility advantage and spatial heterogeneity of successive SARS-CoV-2 variants and their sub-variants circulating between September 2020 and December 2022. Our results show that each variant was progressively more transmissible and more heterogeneously spread. Alpha was 10-40% more transmissible than B.1.177, Delta was 40-100% more transmissible than Alpha, and Omicron was 80-120% more transmissible than the Delta variant. Progressive variants were also more spatially heterogeneous: Alpha was mostly clustered in London and Southeast England, Delta was less clustered and prevalent in both Northwest and Southeast England, while Omicron was dispersed across the country over the first six weeks of transmission. Sub-variants of the same clade didn’t differ in transmissibility or spatial spread. Our method provides a tool for analysis of variant surveillance data, translatable across pathogens and settings, that can capture an emerging variant early and as part of the pandemic preparedness strategy.

## 1 Introduction

Severe acute respiratory syndrome coronavirus 2 (SARS-CoV-2), the virus causing COVID-19, had continued to spread in England throughout 2020, 2021 and 2022. The spread was facilitated by the emergence of new viral variants such as the B.1.177 during the summer 2020, the Alpha and Delta variants, which dominated in late 2020 and early 2021 and the Omicron variants which dominated in 2022. By 30 December 2022, over 20.4 million confirmed cases and over 183 thousand deaths related to COVID-19 had been reported in England [2].

Different non-pharmaceutical interventions (NPIs) including three national lockdowns over 2020 and 2021, with varying levels of social restrictions, Testing-Tracing-Isolation strategies from September 2020, alongside a large scale vaccination strategy from late 2020, have been employed in England to mitigate the epidemic waves caused by these variants. In 2021, alongside the ongoing vaccination against SARS-CoV-2, there was also a gradual lifting of social restriction measures via the reopening Roadmap, starting with reopening of schools in March 2021 as a step 1, and culminating with full lifting of social restrictions from July 2021. In late 2021 and 2022, during the Omicron epidemic waves, additional vaccination strategies were employed including vaccination of adolescents, and boosting immunisation strategies for the elderly.

The emergence of new viral variants, including SARS-CoV-2 variants, is a consequence of mutations of the virus and as a result new variants have potential to be intrinsically more transmissible. Over the last few years, a number of studies have evaluated the transmissibility of different SARS-CoV-2 variants using regression models. We carried out a scoping review of previous findings by searching in Google Scholar for articles containing the words “regression”, “covid”, “variants”, and “transmissibility”. We then checked each result to ensure that it used regression models to quantify the relative transmissibility of variants and discarded any that used other methods, or used regression for other analysis. The estimates for the relative transmissibility of SARS-CoV-2 variants found by the reviewed papers are summarised in Figure 1. Across these studies, the quantified progressive transmissibility of the variants was across different populations, with studies in the USA [8], Japan [17, 27], Norway [13, 15], Spain [29], South Africa [28] and the UK (e.g. [5, 21]). Most studies estimated the transmissibility of variants using case data from population wide testing. However, a few estimated the secondary attack rate (SAR) of the variants, by calculating the number of close contacts of a case who became infected after a given time period, either using contact tracing [15] or by studying the transmission in households and using this to estimate the SAR [3] [13, 17, 27].

**Figure 1.**
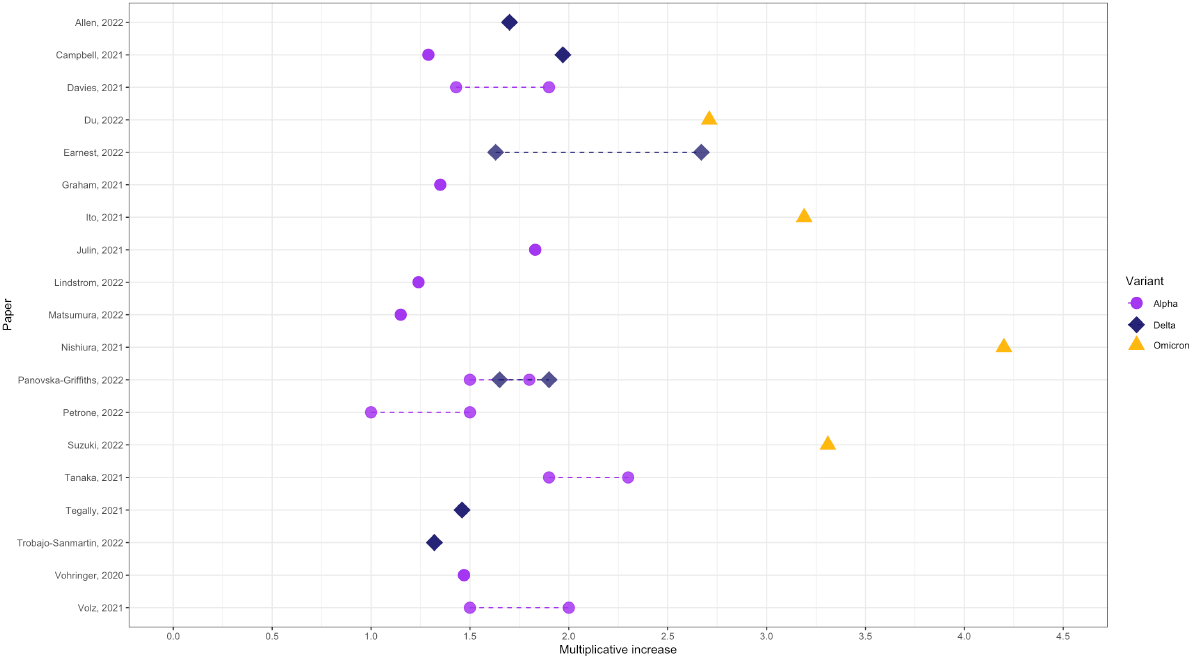
Previous estimates of relative transmissibility of variants of concern from existing published literature. Dashed lines correspond to ranges of plausible values quoted. In each case the multiplicative increase of Omicron is compared to Delta, and Delta is compared to Alpha, apart from in Tegally, 2021, where Delta is compared to Beta. All other advantages are compared to Wild Type or previously dominating variants such as B.1.177. The estimates shown are those stated in the conclusions of the reviewed papers, some gave possible ranges whereas others listed a single estimate.

In the autumn 2020, when the Alpha variant emerged, a number of studies suggested it was more transmissible than the previously dominating B.1.177 variant [4, 5, 11, 13, 15, 17, 21, 23, 27, 30, 31], with estimated advantage ranging from 15% [17] to 130% [27] (circles in Figure 1). The Delta variant, emerging in Spring 2021 and spreading during 2021, was shown across studies to be considerably more transmissible than the Alpha variant [3, 4, 8, 21, 28, 29] (rhombuses in Figure 1). The estimates in increased transmissibility ranged from an increase of 32% [29] to an increase of 167% [8]. Several studies compared the transmissibility of the Omicron variant to that of the Delta variant, [12, 19, 25], with estimates for the relative transmissibility ranging from 3.19-4.20 (triangles in Figure 1). One study, [6], contained a systematic review of studies estimating the reproduction number of the Omicron variant. These authors then compared these estimates to a value for the reproduction number of the Delta variant, calculated in a previous systematic review, [7], and found the pooled estimate for the increase in transmissibility to be 2.71 (with a 95% confidence interval of (1.86, 3.56)). Finally, [16] estimated the reproduction number for variants Alpha, Beta, Gamma, Delta and Omicron, in different countries, using country-wide case data. This study found the highest reproduction number to be that of Omicron, at 1.90 in South Africa, but also found that the estimates for the reproduction number of each variant differed greatly from country to country.

The genetic sequencing of detected cases of SARS-CoV-2 in England by the COVID-19 Genomics UK (COG) consortium has enabled the analysis of the relative properties associated with each variant to be determined. In our previous work we used this data in early 2021, to compare the relative transmission rates of the Delta variant between May and July 2021, to the previously dominating Alpha variant [21]. Here, we extend this work to explore the transmissibility advantage of the progressive variants B.1.177, Alpha, Delta and Omicron variants, and their sub-variants, that were circulating in England between September 2020 and December 2022. We use genetic surveillance data to quantify these but also to compare the progressive spatial heterogeneity over English lower-tier local authority (LTLA) areas. As a sensitivity analysis we also explore whether the progressive transmisibility of the emerging variants changes when we consider fixed or variable variants’ generation time.

## Methods

Using genetic sequencing data from the COG consortium, we developed and applied a set of hierarchical generalised additive models (HGAMs) to each of the successive dominating SARS-CoV-2 variants in England. In the next four subsections we detail the data and the statistical analyses. The models account for variability and structures in the data by allowing smooth functional relationships between predictor(s) and response to vary between groups (such as variant lineage), but in such a way that the different functions are in some sense pooled toward a common shape (such as the overall viral similarities).

### Data

The publicly-available data from the COG consortium [1] corresponded to weekly aggregate counts of each genomic variant sequenced at the Wellcome Sanger Institute to monitor COVID-19 dynamics within each LTLA of England. The dates correspond to the date on which the sample was collected.

The corresponding latitude and longitude of each LTLA were obtained from [10] and matched to the LTLA code in the COG data. These were then used to inform the spatial variation in counts within the regression model. The temporal aspect of the model is informed by the number of weeks since the start of the study period (here September 5th 2020). The raw data are shown in Figure 2. We also use vaccination data from NHS England’s weekly vaccination reports [9] to account for impacts of vaccination on the observed growth rates of emerging variants.

**Figure 2.**
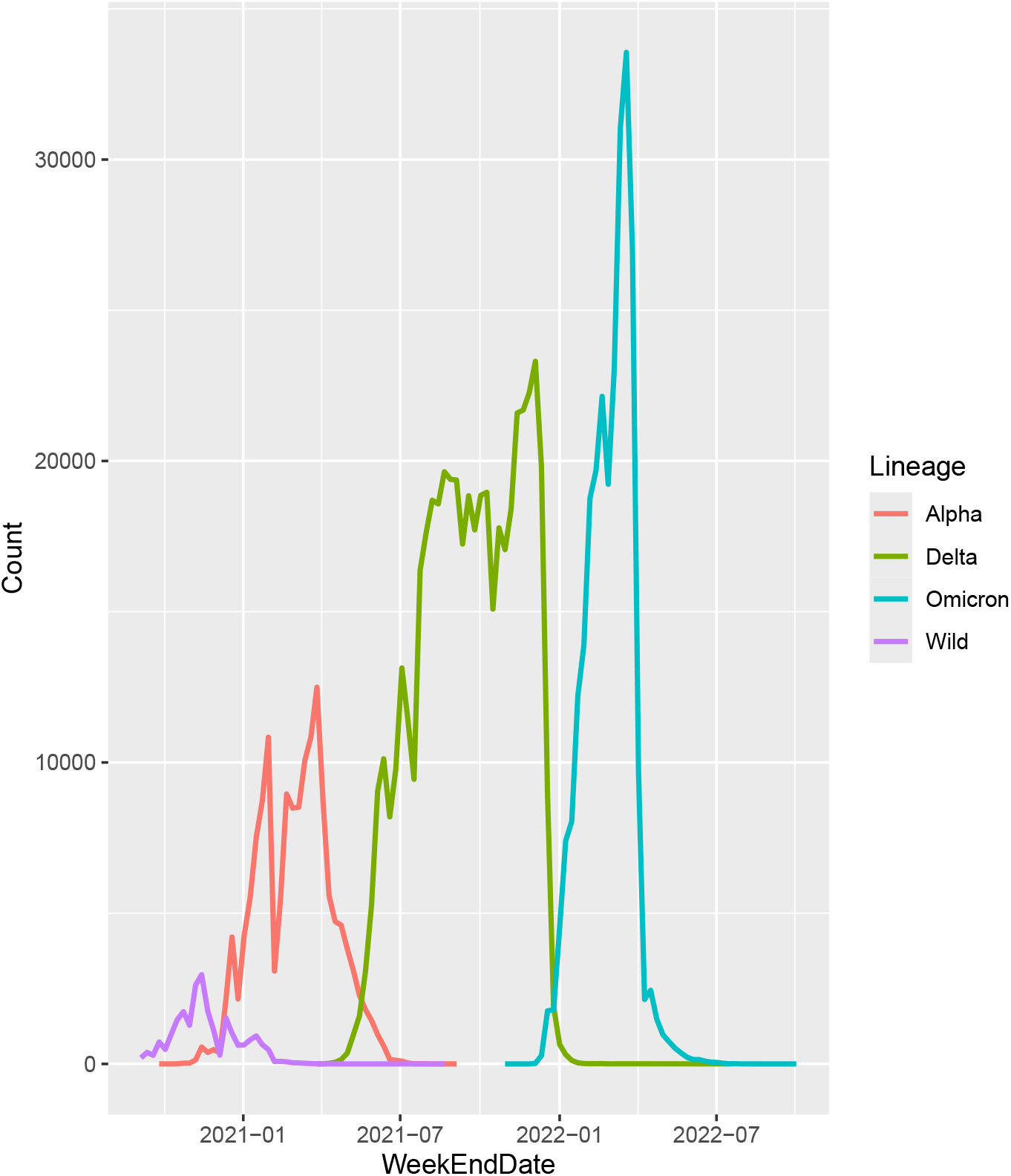
Raw data showing observed counts of the successive principal variants sequenced through time. Data from COG consortium [1].

### Statistical Models

We adapt and extend methodology proposed in [21] and [5], to determine the relative multiplicative rate of transmission of competing variants and subvariants across LTLAs in England. Hierarchical generalised additive models (HGAMs) [22] with a negative binomial response distribution were fitted with count of each variant per week and per LTLA as the response, and using a GI structure as described in [22]. The mean structure is modelled as a function of fixed effects and smooths of day, latitude, longitude and lineage. For each smooth, the form of the basis function induces a penalty matrix **S**, which is pre- and post-multiplied by the parameter vector *β* and a smoothing parameter *λ*. With grouped data, there is a choice in how the smoothing should be carried out and what level in the hierarchical structure. In [21], the optimal models were shown to be those with a global smoother plus group-level smoothers with differing degrees of smoothing (the GI model). In this model, the mean structure is modelled as follows:

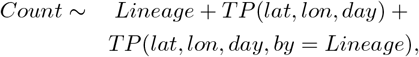

where the Lineage effect provides a intercept baseline for each variant and *TP* is a tensor product smooth of thin plate splines. This contained both a global thin plate regression spline and thin plate regression splines fitted separately to each lineage, in which the parameters and smoothness were allowed to vary between lineages. These allow for heterogeneity in observed counts across variants, across space and time and through a penalised smoothed combination of the two. This will represent differences in inherent epidemiology of the variant (fixed intercept), seeding location (space-time smooth) and any residual combination of the two (space-time-lineage smooth). The dimensions of the bases were assumed to be 10 for contiuous variables and 6 for random effects, with spline order two for each smooth function. The `gam.check` function from the `mgcv` package was used to ensure appropriate number of effective degrees of freedom and knot locations had been used. Four simpler models were also fitted in each case but were discarded due to higher Akaike Information Criteria (AIC). Model residuals can be checked for patterns across covariates marginally in both one- and two-dimensions. Strong patterns would suggest important components in the data the model is not well capturing and as the approach is aiming to interpolate the data, out of sample prediction is not required.

### Quantifying progressive variants’ transmissibility

We initially studied the relative transmissibility of combined principal lineages relative to the previous, where the count of cases was summed over principal sublineages of the variant. Conditional on the fitted model, each LTLA was treated as a separate experimental unit, which may be correlated to a lesser or greater degree to other LTLAs. Relative multiplicative transmission rates were estimated for each LTLA using linear trends extrapolated from the fitted hierarchical models, as interactions across the determined dates of emergence. Linear slopes during the emergence period (Table 1) for each (sub)variant were calculated using the `emmeans` package [14] and the ratios of these slopes determined in each LTLA.

**Table 1.** Table showing the period of interest for each variant derived from the emergence periods for each variant according to data from the COG consortium (author?) [1].

| Variant | Date Range |
| --- | --- |
| B.1.1.7 | 24/10/2020 - 02/01/2021 |
| B.1.617.2 | 05/05/2021 - 17/07/2021 |
| AY.4 | 05/05/2021 - 17/07/2021 |
| BA.1.1 | 13/12/2021 - 07/02/2022 |
| BA.2 | 17/01/2022 - 21/03/2022 |

This way we compared relative rates of transmission of B.1.177 vs Alpha, Alpha vs Delta and Delta vs Omicron variants i.e. each successive variant relative to the previously circulating variant over the time period in which they showed exponential growth across England. We then compared the Delta and Omicron sub-variants to the previously dominating sub-variant. Specifically, we analysed the transmissibility of sub-variants: B.1.617.2 vs the Alpha variant, the B.1.617.2 and Delta AY.4 variant, the Delta A4.4 variant and the Omicron BA1.1 variant, and the Omicron BA.1.1 and BA.2 variants; these were circulating over 2021 and 2022 period for which the COG data was available. The time period of interest for each variant and sub-variant is shown in Table 1. We note that for the comparisons using combinations of variants (i.e. Alpha vs Delta and Delta vs Omicron), we used the union of the time periods of the emerging sub-variants involved.

Unlike in [21], we fitted the model to the sub-variants pairwise, and so the values for the relative multiplicative rate of transmission depended only on the number of cases of the two sub-variants being compared. This meant that the calculation for the relative multiplicative advantage was not influenced by previous variants affecting the global smoother. Additionally, the multiplicative outcome can in this way be proxied by a multiplicative temporal reproduction number *R*_*t*_, which gives the secondary number of cases emerging from an infected case. We also compare the estimated linear trends against lagged measures of seropositivity in the same spatial region using Pearson’s correlation coefficients.

### Correlating transmissibility to vaccination and seropositivity

We investigated the association between relative transmissibility and vaccination uptake (first, second, and third doses) across LTLAs. Weekly vaccination levels from two months prior to variant emergence up to the first week included in the transmissibility estimates were compared with the relative transmissibility using Pearson’s correlation coefficients 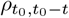, for *t* ∈ 8, …, 0, where *t*_0_ is the first week from Table 1.

To explore the relationship between transmissibility and vaccination, we carried out a Pearson’s correlation test comparing the proportion of the population in each LTLA who had been vaccinated with the estimates for the relative transmissibility of each variant in that region. To account for any delay in reporting vaccination numbers, and time to build immunity, we estimated the correlation using vaccination numbers from each of the 10 weeks prior to the emergence of the new variant. The vaccination roll-out had not begun at the time of the Alpha variant’s emergence, therefore we were only able to investigate the correlation for the relative transmissibility of Delta over Alpha, and Omicron over Delta. We also carried out the same analysis for the relative transmissibility of the sub-variants B.1.671.2 over Alpha, and BA.1.1 over AY.4.

Data on seropositivity were taken from the Office for National Statistics (ONS)[20]. Weekly measurements of seropositivity (179 ng/ml antibody level across England) were averaged across age classes. The total number of positive tests in a given week were summed over all spatial regions to give a comparable resolution to the seropositivity data. The response variable was taken to be the total weekly counts of positive PCR tests for each lineage, and this was matched to the corresponding nearest seropositivity measurement in time, conditional on that measurement being no later than the PCR total count. The generalised additive model with negative binomial response distribution was constructed with a linear predictor consisting of lineage-specific intercept, independent smooths of day and seropositivity, and a random effect tensor product thin-plate spline of seropositivity and day (10 knots each), with the random effect being lineage. The model was again checked for convergence and sufficient flexibility in knot numbers and location.

### Quantifying the spatial heterogeneity in successive variants

The estimated relative transmissibilities from each of the analyses were then tested for spatial auto-correlation using Moran’s I statistic [18]. The statistical test determined evidence against the null hypothesis of no spatial auto-correlation, which would correspond to random spread across LTLAs. Higher positive test statistics tended towards significant p-values and indicated local clustering of high transmission during the period of emergence, whilst significant negative values indicated dispersion or anti-correlation.

## Results

### Statistical models fit the data

The GI models outlined above were fitted to successive (sub-)variants of SARS-CoV-2. Model checks were carried to ensure sufficient flexibility and numbers of knots were included. Residual plots across latitude, longitude and day and across latitude and longitude were generated (Supporting Information) to check for patterns that would indicate poor model fit. All checks suggested a well-fitted and calibrated statistical model.

### Quantifying the progressive transmissibility advantage of successive overall Alpha, Delta and Omicron variants

Our results showed that each new emerging variant was more transmissible than the previously dominant variant. Alpha was 10-40% more transmissible than B.1.177 (Figure 3a), Delta was 40-100% more transmissible than Alpha (Figure 3b), and Omicron was 80-120% more transmissible than the Delta variant (Figure 3c).

**Figure 3.**
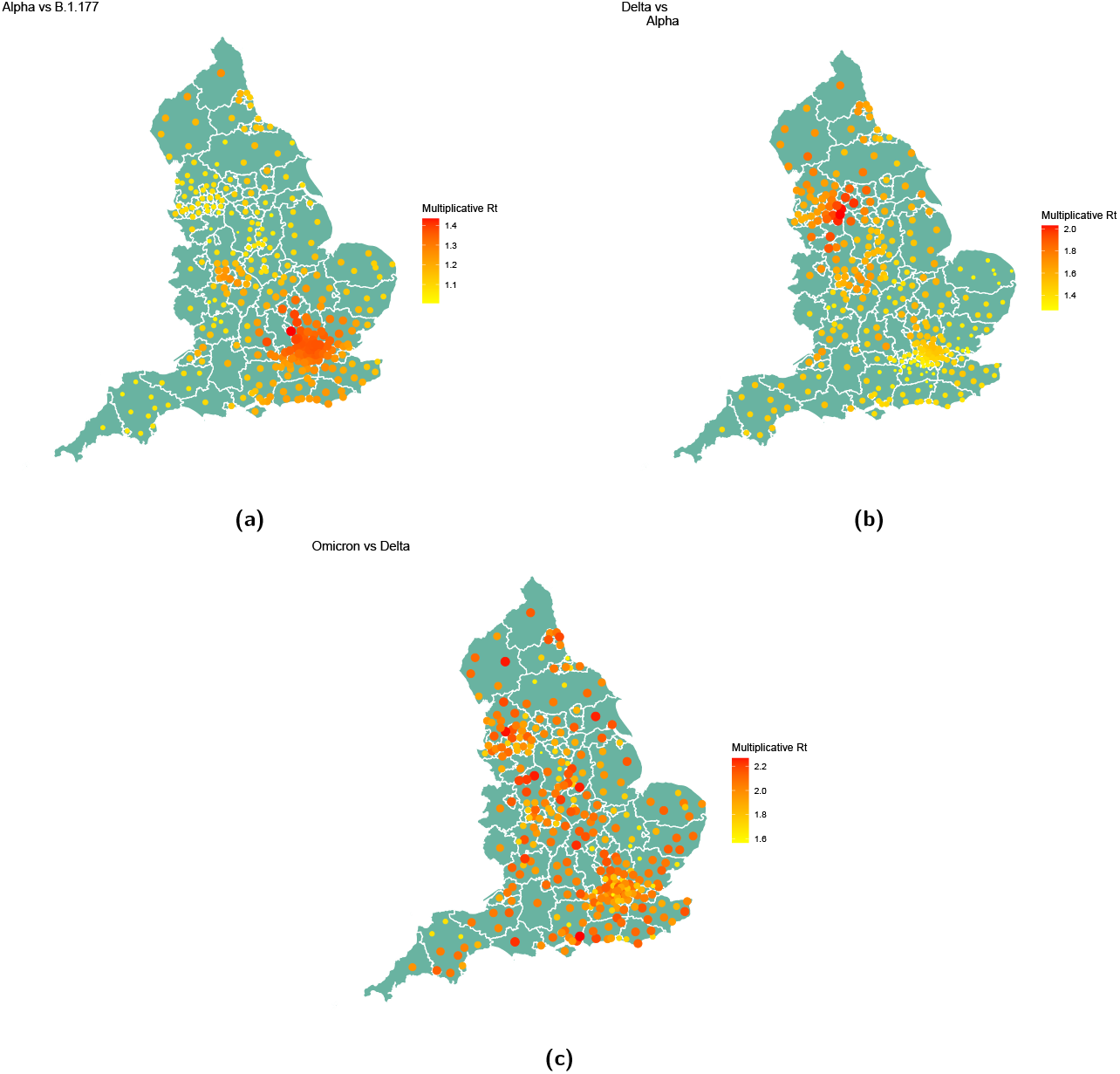
Maps showing progressive transmissibility advantage of the Alpha, Delta and Omicron variants across LTLAs in England.

### Quantifying the progressive transmissibility advantage of emerging sub-variants

When comparing sub-variants, we found large variability in relative transmissibility of successive sub-variants. Successive sub-variants that belonged to a principal variant different to the one previously dominating, were always more transmissible. For example, the B.1.617.2 variant was 10-40% more transmissible than the previous Alpha variant (which didn’t have sub-variants), while the Omicron BA.1.1 variant was 20-60% more transmissible than the previously dominating Delta AY.4 variant (Figure 4c). However, when the emerging sub-variant belonged to the same principal variant (such as the Delta B.1.617.2 and AY.4 variants or the Omicron BA.1.1 and BA.2 variants), we didn’t find significant difference in their progressive transmissibility (Figure 4).

**Figure 4.**
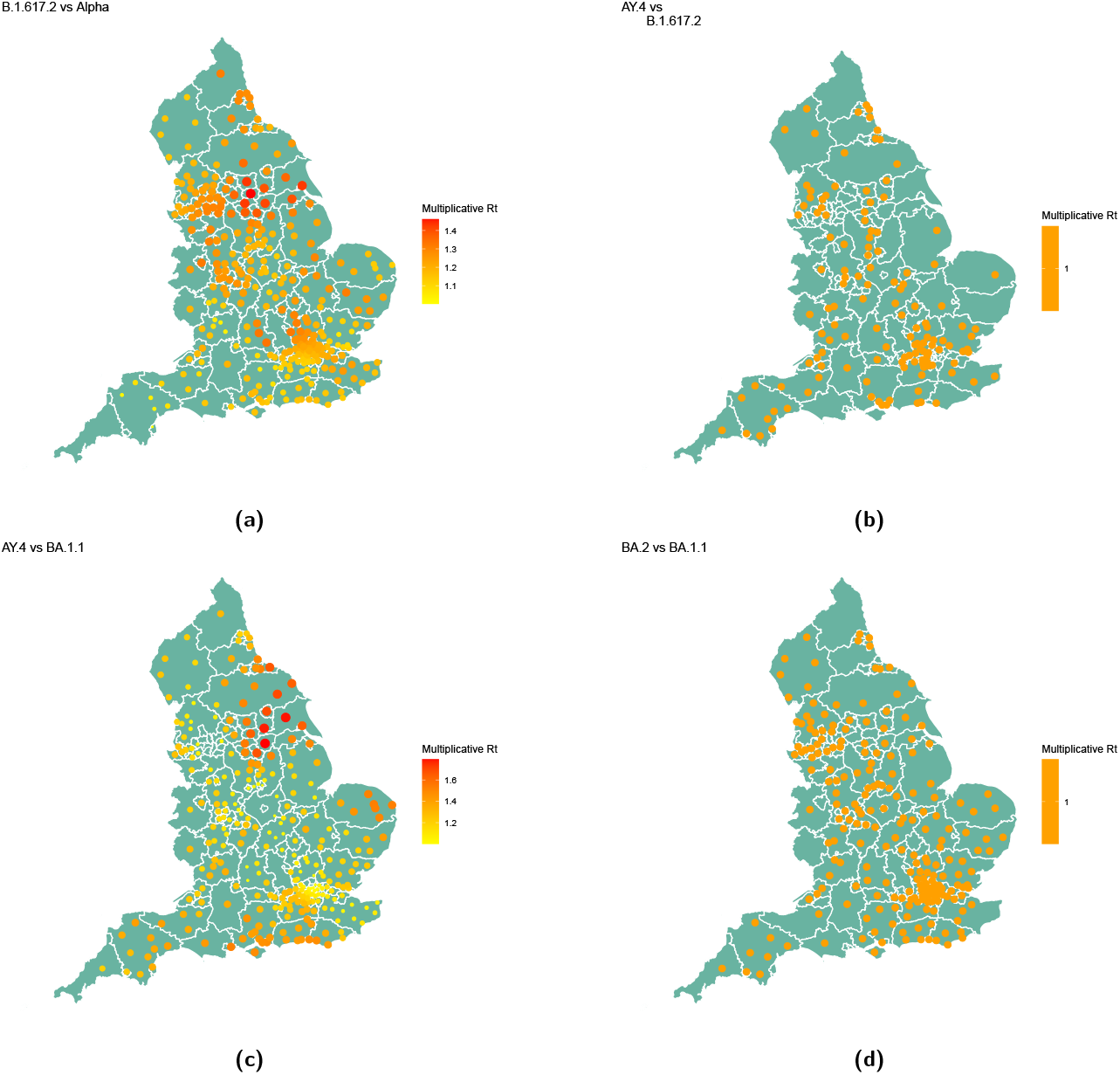
Maps showing the progressive transmissibility advantage of each sub-variant.

### Spatial heterogeneity and clustering

The Moran’s I statistics (Table 2) suggested significant clustering (positive test statistics) across all variants, but no significant spatial structure within variants. For example, progressive variants were more spatially heterogeneous over the first six weeks of transmission: Alpha was mostly clustered in London and Southeast England (Figure 3a and Table 2), Delta was less clustered and prevalent in both Northwest England and Southeast England (Figure 3b and Table 2), while Omicron was heterogeneously spread across the country and not as evidently clustered (Figure 3c and Table 2). Emerging sub-variants were more clustered if they belonged to different principal variant. For example, both the Delta 1.617.2 sub-variant, which emerged after the Alpha variant, and the Omicron BA.1.1 variant, which followed the AY.4 Delta variant, were more clustered around London and across Northeast England (Figure 4). The Delta AY.4 variant, which followed the Delta B.1.617.2 variant, was more heterogeneously spread, as was the Omicron BA.2 compared to the Omicron BA.1.1 variant.

**Table 2.** Moran’s I statistics and additional hypothesis test summaries for estimates of relative transmissibility between successive variants and sub-variants. P-values reported to 3d.p.

| Comparison | Observed MI | Sd | P-value |
| --- | --- | --- | --- |
| Alpha vs B.1.177 | 0.418 | 0.005 | 0 |
| Delta vs Alpha | 0.325 | 0.005 | 0 |
| Omicron vs Delta | 0.279 | 0.005 | 0 |
| B.1.617.2 vs Alpha | 0.170 | 0.005 | 0 |
| AY.4 vs B.1.617.2 | -0.002 | 0.005 | 0.824 |
| BA.1.1 vs AY.4 | 0.158 | 0.005 | 0 |
| BA.2 vs BA.1.1 | -0.008 | 0.0050 | 0.412 |

#### Effects of variable generation time

When we considered the emerging variants to have variable generation time, our results remained the same. Namely, each new emerging variant was again more transmissible than the previously dominant variant. However, the relative progressive transmissibility difference between successive variants was less when generation time was variable, than when it was fixed. Delta was up to 10% more transmissible than Alpha (SI Figure 3), and Omicron was 5% more transmissible than the Delta variant (SI Figure 4).

Similarly each progressive variant was spatially more heterogeneous and more wide spread than the previous circulating variant. The magnitude of difference, however, was lower, suggesting some of the initial gain of novel variants was due to a reduced generation time (Supporting Information Figures 3-4).

#### Effects of vaccination

Vaccination was generally negatively correlated, at 5% significance level, with transmissibility across the variants. Our results suggest that, at the 5% significance level, the proportion of the population vaccinated with first and second doses was negatively correlated with the relative transmissibility of Omicron over Delta, with Pearson’s correlation coefficients varying between −0.118 and −0.126 for first doses, and between −0.114 and −0.121 for second doses, depending on the week used. There was also evidence, at the 5% significance level, to suggest that the vaccination level for first and second doses was negatively correlated with the relative transmissibility of B.1.617.2 over Alpha, with Pearson’s correlation coefficients varying between −0.150 and −0.185 for first doses, and between −0.123 and −0.149 for second doses, depending on the week used.

Predicted counts of positive tests across seropositivity and each lineage are shown in Figure 5. The lineage fixed effect and smooth of seropositivity were weakly significant at the *α* = 0.1-level (SI Table 6), with the fitted smooth term for seropositivity on the link-scale shown in Figure 5. The overall effect is marginally negative. The factor-level intercept increases with each emerging variant (i.e., highest for BA.1.1). Estimated over-dispersion parameter for the response distribution is *θ* = 13.567, supporting the use of a negative binomial over a Poisson model.

**Figure 5.**
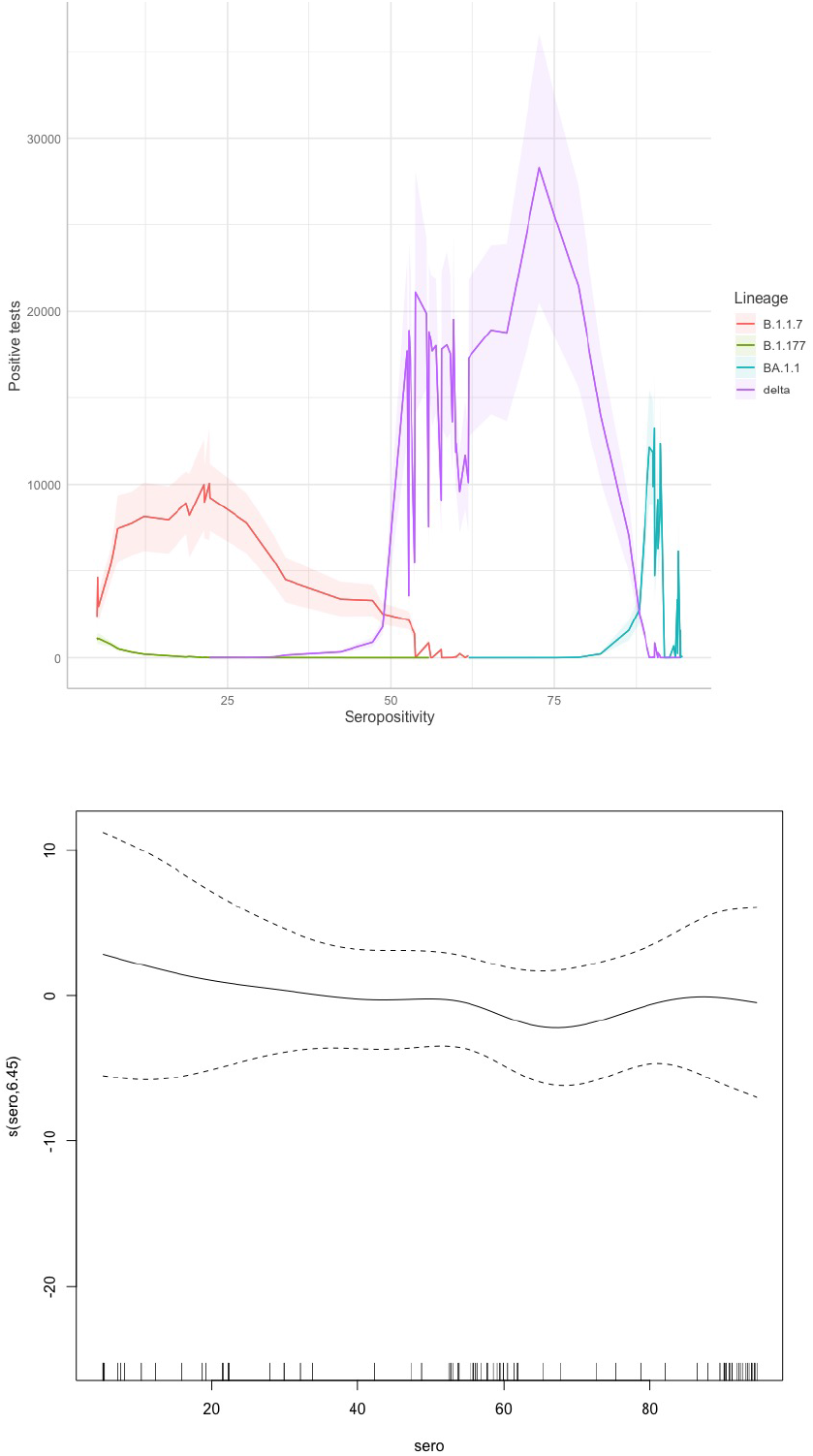
Predicted counts of positive tests across observed seropositivity (upper) and estimated smooth effect of seropositivity, ‘s(sero)’, on the scale of the linear predictor (lower).

## Discussion

We use genetic surveillance data and flexible regression models to quantify the progressive advantage and spatial spread of SARS-CoV-2 variants and sub-variants that were circulating in England over periods between September 2020 to December 2022. Our results suggest that emerging variants were progressively more transmissible and more heterogeneously spread across England: Alpha was 10-40% more transmissible than B.1.177, Delta was 40-100% more transmissible than Alpha, and Omicron was 80-120% more transmissible than the Delta variant. Progressive variants were also more widely spread over the first six weeks of transmission: Alpha was mostly clustered in London and Southeast England, Delta was less clustered and prevalent in both Northwest England and Southeast England, while Omicron was heterogeneously spread across the country. Finally, we show that successive sub-variants that belonged to the same principle variant were more transmissible and spatially spread than the previous ones (e.g. the Omicron BA.1.1 variant was 20-60% more transmissible than the previous Delta AY.4 variant). This was not the case with emerging sub-variants from the same principal variant, with our results showing that the consecutive Delta and Omicron variants had similar transmissibility and spatial spread. Modelling variable generation time didn’t affect our findings, but when variable generation times were incorporated into the modelling of successive variant transmissibility, the relative difference in their progressive superiority was less.

The distribution of multiplicative rates is bimodal in nature. Around one third of the LTLAs observe a relative *R*_*t*_ close to one, suggesting that the variant does not out-compete the other circulating variants. For those LTLAs in which the new variant does become a dominant variant, there is a relatively symmetrical distribution of multiplicative rates.

Our results of progressive transmissibility of successive principal variants are in line with previous findings. As noted in our scoping review of previous results in the introduction, Alpha was 15-130% more transmissible than the B.1.177 variant, with our determined range well within this. Similarly, Delta was found in previous studies to be 32-167% more transmissible than the Alpha variant, and this is similar to our range of 40-100% advantage in transmissibility. A smaller number of studies have previously quantified the transmissibility advantage of Omicron vs Delta to be between 80-320%, and this range includes our predicted range of 80-120%. Overall, our results add to the existing literature. But importantly, unlike existing studies, as well as comparing principle variants, we also compared successive sub-variants. Our results that transmissibility advantage is a characteristic of sub-variants from a new principal variant, are novel. Furthermore, we also showed that successive new variants, and sub-variants of different principle variant, were more spatially spread and less clustered. Overall, we found that with each emerging variant, whilst still statistically significant, the level of clustering determined by the Moran’s I test statistic has reduced. This implies a tendency towards more random spread of the disease with genetic evolution of variants. Both of these results are useful in preparing surveillance and response when a new principal variant, or a new sub-variant emerges.

It is important to note that as progressive COVID-19 variants emerged, different public health interventions were also rolled out, and the population started to get immunised or infected or both. Hence the degree to which the variant could evade immunity, in addition to the variant’s transmissibility, could be relevant. To address this, we investigated whether the relative transmissibility of variants was affected by the roll out of vaccinations across England and also whether seropositivity was associated with increased transmissibility.

Given the sequential nature of the variants and the lack of ability to experimentally allocate seropositivity to different lineages, the estimated association between transmissibility and seropositivity is mixed. There is weak significance of lineage and seropositivity, suggesting the lineage is the more important aspect for determining transmissibility, with a small overall effect of prior immunity on reducing the transmission further. The lack of strong statistical significance in the GAM terms could stem from the aggregation over spatial regions, preventing signal from spatial heterogeneity to be utilised. Alternatively, the individual sequential nature of immune response in the virus evolution could explain the lack of strong dependence on this prior infection. This is particularly evident in the Delta wave, where higher seropositivity is associated with a higher number of positive tests. These results suggest it is difficult to say that anything but progressive transmissibility and spatial heterogeneity is crucial for ongoing emergence of COVID-19 variants.

Our study has a number of strengths. Firstly, we extended and adapted established and existing methodology to longer time periods and sub-variants. We evidence the important observation that if we have groups of variants (e.g. all of Delta or all of Omicron) we have enough power to detect differences, while if we look at sub-variants it is more difficult to get significant results. Whether this is due to a weaker signal from shorter circulating times and greater numbers of sub-variants circulating, or a closer genetic similarity in those sub-variants, is less clear. Using flexible regression models, compared to other published studies, allows analysis of large data sets, including spatial smoothing and predictive ability, which can be achieved with good surveillance.

Overall, using flexible statistical models, we were able to show that successive variants of SARS-CoV-2 in England between September 2020 and December 2022 were progressively more transmissible and increasingly more widely spread across England. Importantly, genetically new principal variants or sub-variants of different clade, have a tendency to be more intrinsically transmissible and able to spread widely. This highlights the importance of continued surveillance to capture an emerging variant early, as this will allow for timely local and targeted interventions to prevent wider and larger-scale spread of emerging variants and improved pandemic preparedness.

Our study is the first to consider both temporal and spatial heterogeneity of emerging SARS-CoV-2 variants using genetic data. Our findings of spatial homogeneity/heterogeneity and clustering estimates support a tendency of new and more mutated variants to spread more rapidly and over larger space. Spatial modelling of adaptive viruses remains a significant challenge in [24, 26]. The advantage of regression-based statistical models is the ability to incorporate additional features or factors into the modelling framework to explain the variability in spatial spread and improve predictive ability of the models.

Whilst these are not formal experimental conditions, they do enable the comparison of relative transmission rates observed in the wider population as if they were an experimental study. Similarly, LTLAs in England, assuming that any underlying correlation between them has been accounted for, can also be treated as multiple experimental units. Determining the relative behaviours of the transmission rates across the population and between different variants that emerged has the potential to highlight important variation in transmission across space and between different successive variants and against the wild-type SARS-CoV-2 variant. The results support previous analyses from more directed methods for detecting relative transmissibility of emerging variants of concern.

One challenge of using happenstance data is that there is a restriction to utilise the data as they have been collected. In this instance, it was not possible to look at the course of the entire epidemic since the genetic data did not have enough power for the later Omicron variants when surveillance was reduced. Combining different datasets as proxy for different variants is an alternative way to amplify the size of our datasets. For example, we could use S gene dropout or S gene target failure (SGTF) data, which we refer to if the commercial SARS-CoV-2 PCR test is unable to detect the S gene target. Over the COVID-19 epidemic, the SGTF signature has been used as a proxy to preliminarily infer the presence of the Alpha and Omicron variants of SARS-CoV-2. Putting this in the context of our study, we could use SGTF data from PCR testing from the periods we studied as a proxy to amplify the size of the Alpha and Omicron variants. But this would imply that we are using a different data set to that for the Delta variant. We note that mixing data types is not guaranteed to improve inference or prediction. To do this properly, much consideration would need to be given to how the two data types are combined and how they would interact in the statistical observation model. This is far from trivial given that multiple variants are circulating simultaneously, so SGTF is not a 1-to-1 relationship with a given (sub)-lineage. Furthermore, employing this in our models will mean that we will be comparing different datasets with the same models and inferring results across different datasets. Performing this requires special care and new statistical methodologies to ensure appropriate calibration. Using and linking different datasets is not something we have set out to do, and we also don’t have access to all the datasets to do this precisely. This is something we feel we can explore in future, but it is out of scope in this paper. Instead we note that continued and sustained surveillance is therefore crucial if methods like this are used as a public health benefit.

In our analysis we have used the relative exponential growth rates of the sequence counts of variants as a measure for comparing the growth advantage of different variants. An alternative metric to this would be the growth rate of the relative frequency of a new variant among all sequenced samples – as an estimate of that variant’s relative fitness (or selective advantage) compared to other co-circulating variants. The metric we used gives the exponential increase in the frequency (or counts) of a particular genetic sequence (e.g., a new viral variant) within a collected set of sequence data over time. For the purposes of our analysis in this study, we wanted to quantify the rapid increase in frequency in the dataset to mirror the real-world biological spread and transmission advantage of that variant in a population, and importantly compared to a previous variant and hence we chose to use the relative exponential growth rates of the sequence counts of variants. The core principle of understanding is that variants with higher relative fitness values will increase in frequency within the population at an exponential rate relative to other, less fit variants. Hence, in theory there should be a direct correlation between these two metrics, as greater positive relative fitness directly corresponds to a steeper, faster exponential growth in the observed count in that variant’s sequence data in the genomic surveillance. Since we were using genetic data, the latter metric was more sensible to use for us.

From the definition of these two metrics, we appreciate that the growth rate of the relative frequency of a new variant among all sequenced samples can comprise competitive advantage, replacement dynamics and public-health-indicators related to the variant. Therefore it could be argued that this may be considered a more robust measure of the true growth advantage of that variant than a comparison of the relative exponential growth rates of the sequence counts of variants. However, using the frequency metric can be heavily influenced by changes in testing capacity, sequencing efforts, and population behaviour, especially as mandatory public health and social measures were different across different variants. This makes it more difficult to make direct comparison across different time periods or regions when using the growth rate of the relative frequency of a new variant among all sequenced samples as a metric. This is why for the purposes of the study, and within the scope to compare relative variants’ transmissibility over time and regions, we feel that using the relative exponential growth rates of the sequence counts of variants is a sufficient measure when comparing growth advantage of different variants.

We showed there was substantial heterogeneity in transmissibility between sub-variants of different lineages, but there was a lack of power in detecting significant differences in transmissibility between sub-variants of the same dominant variant. Additional challenges were observed in later sub-variants of Omicron as wide-scale national testing was terminated. Our results suggest that this is indeed the case for all Omicron sub-variants were more transmissible than AY.4. Figure 4d also shows the spatial variability in relative transmissibility for BA.1.1 and BA.2 relative to delta variant AY.4.

From publicly available data, we were able to replicate more labour and cost-intensive methods for estimating transmissibility. In addition our analyses are able to determine spatial variability in observed transmissibility. One of the challenges in these observed data are separating out the impacts of changes in pharmaceutical and non-pharmaceutical interventions from the inherent variation in the transmissibility of the variants. The upper bound on ranges in relative transmissibility frequently correlates well with previous more direct estimates of transmissibility, in areas where interventions are perhaps not having significant impact, however there are frequently regions where the transmissibility is coupled with interventions that impact transmission where the observed transmission may not reach its full potential.

In summary, in this paper, we show how combining robust statistical models with genetic data can be used to quantify the transmissibility and dispersion advantage of emerging pathogens, here proxied by different SARS-CoV-2 variants and sub-variants. Hence we illustrate how this can be readily used as part of an improved pathogen surveillance and pandemic preparedness strategy.

## Author contributions statement

BS developed the methodology and wrote the initial computational code with input from JPG. JG and BS ran the analyses and produced the Tables and Figures. All authors contributed to the writing of the first draft of the manuscript. All authors provided critical feedback and helped shape the research, analysis and manuscript. All authors have approved the manuscript and gave their consent for submission and publication.

## Data availability

Data and code to reproduce the analyses and figures presented in the paper are available at https://github.com/joshuagrier/COGSpatial-transmissibility.

