## Supplemental Information for "Spatio-temporal transmissibility and dispersion of SARS-CoV-2 variants and sub-variants of concern in England"

### 1 Appendix A

#### Methods

We fitted the equivalent models from [Pedersen et al., 2019] for our data. That is:

1. Model G - the smoother is fixed across all variants.
2. Model S - the smoother is estimated for each variant separately but each smoother has the same wiggliness (smoothness).
3. Model I - the smoother is estimated for each variant separately with different wiggliness.
4. Model GS - a global smoother is added to model S.
5. Model GI - a global smoother is added to model I.

The models are constructed in the `mgcv` package as follows.

```
cov_modS <- bam(Count ~ t2(lat, lon, day, Lineage, bs=c("tp", "tp", "tp", "re"),
                           k=c(10, 10, 10, 6), m=2, full=TRUE),
                data=d2, method="fREML",discrete=T, family="nb",nthreads=3)
```

The function `t2` defines the 2-dimensional smoothers for the model. The first terms listed inside `t2` are the variables which the smoother will be a function of. `bs` defines the type of basis function which will be fitted for each variable, in this case `tp` is a thin plate regression spline and `re` is a penalised parametric randome effect term. `k` specifies the knots for each of the smoother terms and `m` defines the order of all the splines. We specified the response

[1]School of Mathematics and Statistics, Center for Research into Ecological and Environmental Modelling, University of St Andrews, UK
[2]University of Oxford, OX1 4BH, Oxford, UK
[3]The Queen's College, University of Oxford, OX1 4BH, Oxford, UK
[3]Pandemic Sciences Institute, University of Oxford, OX3 7LF, Oxford, UK

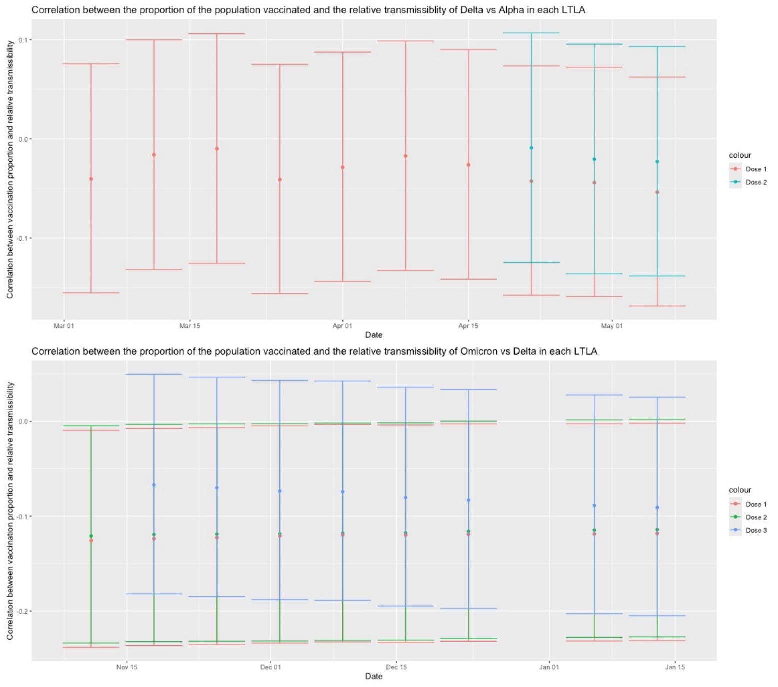


Figure 1: Plots showing the correlation between the relative transmissibility of the variants and the proportion of the population who had received each vaccine dose in each LTLA. The correlation shown in estimated by Pearson's Correlation Coefficient.

distribution as negative binomial and this was the same for all of the models used (in the code the distribution is specified by `family="nb"`).

Model I is very similar, coded as follows:

```
cov_modI <- bam(Count ~ Lineage + te(lat, lon, day, by=Lineage,
                                     bs=c("tp", "tp", "tp"), k=c(10, 10, 10), m=2),
                data=d2, method="fREML", family="nb",nthreads=3)
```

The main difference here (apart from the use of the `te` smoother instead of `t2`) is that we have allowed the smoother to have a different parameter in each group by fitting to lineage outside of `te` and by using the `by` argument on the lineage groupings.

For model G a global smoother, which did not depend on lineage, was fitted using the `t2` function:

```
cov_modG <- bam(Count ~ t2(lat, lon, day, bs=c("tp", "tp", "tp"), k=c(10, 10, 10)),
                data=d2, method="fREML",discrete=T, family="nb",nthreads=3)
```

For models GS and GI, the global smoother specified in G was added to models S and I respectively.

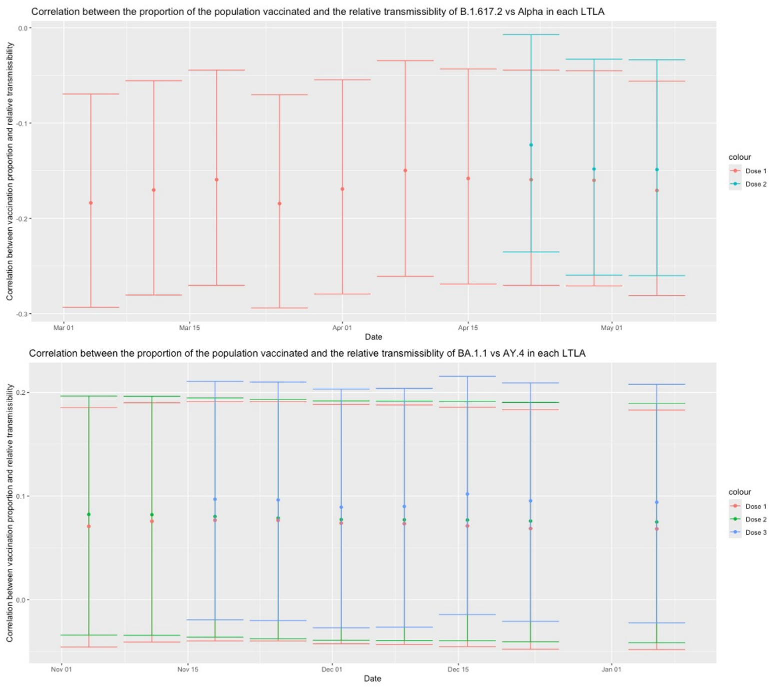


Figure 2: Plots showing the correlation between the relative transmissibility of the sub-variants and the proportion of the population who had received each vaccine dose in each LTLA. The correlation shown in estimated by Pearson's Correlation Coefficient.

Table 1: Table showing the p-values for the correlation estimates between the proportion of the population vaccinated and the relative transmissibility of different SARS-CoV-2 variants in each LTLA. The lag (in weeks) is the number of weeks prior to the emergence of the new variant emerging at which the vaccination data is used.

| | | p-value | | | | | | | | | |
|---|---|---|---|---|---|---|---|---|---|---|---|
| Lag (weeks) | | 0 | 1 | 2 | 3 | 4 | 5 | 6 | 7 | 8 | 9 |
| Delta vs Alpha | Dose 1 | 0.364 | 0.456 | 0.472 | 0.659 | 0.771 | 0.630 | 0.489 | 0.867 | 0.786 | 0.496 |
| | Dose 2 | 0.699 | 0.729 | 0.878 | - | - | - | - | - | - | - |
| | Dose 3 | - | - | - | - | - | - | - | - | - | - |
| Omicron vs Delta | Dose 1 | 0.046 | 0.052 | - | 0.045 | 0.043 | 0.044 | 0.041 | 0.038 | 0.037 | 0.034 |
| | Dose 2 | 0.054 | 0.053 | - | 0.050 | 0.047 | 0.047 | 0.045 | 0.045 | 0.044 | 0.042 |
| | Dose 3 | 0.126 | 0.135 | - | 0.161 | 0.176 | 0.212 | 0.217 | 0.238 | 0.260 | - |
| B.1.617.2 vs Alpha | Dose 1 | 0.004 | 0.007 | 0.007 | 0.007 | 0.011 | 0.004 | 0.002 | 0.007 | 0.004 | 0.002 |
| | Dose 2 | 0.012 | 0.012 | 0.037 | - | - | - | - | - | - | - |
| | Dose 3 | - | - | - | - | - | - | - | - | - | - |
| BA.1.1 vs AY.4 | Dose 1 | 0.250 | - | 0.248 | 0.231 | 0.217 | 0.214 | 0.197 | 0.197 | 0.203 | 0.234 |
| | Dose 2 | 0.207 | - | 0.202 | 0.196 | 0.195 | 0.193 | 0.185 | 0.177 | 0.167 | 0.166 |
| | Dose 3 | 0.113 | - | 0.108 | 0.086 | 0.130 | 0.133 | 0.105 | 0.102 | - | - |

Table 2: Table showing estimates for relative increase in transmissibilities of different variants, with 95% CIs shown in brackets where they are stated in the literature.

| Paper | Alpha | Beta | Gamma | Delta | Omicron |
|---|---|---|---|---|---|
| Allen et al. [2022] | - | - | - | 1.70 | - |
| Campbell et al. [2021] | 1.29 (1.24-1.33) | 1.25 (1.20-1.30) | 1.38 (1.29-1.48) | 1.97 (1.76-2.17) | - |
| Davies et al. [2021] | 1.43-1.90 (1.38-2.30) | - | - | - | - |
| Du et al. [2022] | - | - | - | - | 2.71 (1.86, 3.56) |
| Earnest et al. [2022] | - | - | - | 1.63-2.67 | - |
| Graham et al. [2021] | 1.35 (1.02-1.69) | - | - | - | - |
| Ito et al. [2022] | - | - | - | - | 3.19 (2.82-3.61) |
| Julin et al. [2021] | 1.83 (0.86-3.22) | - | - | - | - |
| Lindstrøm et al. [2022] | 1.24 (1.20-2.14) | - | - | - | - |
| Matsumura et al. [2022] | 1.15 | - | - | - | - |
| Nishiura et al. [2021] | - | - | - | - | 4.20 (2.10–9.10) |
| Panovska-Griffiths et al. [2022] | 1.50-1.80 | - | - | 1.65-1.90 | - |
| Petrone et al. [2022] | 1.00-1.50 (0.75–1.71) | - | - | - | - |
| Suzuki et al. [2022] | - | - | - | - | 3.31 (2.95–3.72) |
| Tanaka et al. [2021] | 1.90-2.30 (1.47-3.21) | - | - | - | - |
| Tegally et al. [2021] | - | - | - | 1.46 (1.44-1.48) | - |
| Trobajo-Sanmartín et al. [2022] | - | - | - | 1.32 (1.13-1.53) | - |
| Vöhringer et al. [2020] | 1.47 (1.41-1.53) | - | - | - | - |
| Volz et al. [2021] | 1.50-2.00 (0.09-2.65) | - | - | - | - |

The Omicron transmissibility is compared to that of Delta, and in each case Delta is compared to Alpha, apart from in Tegally et al. [2021], where it is compared to Beta, all other comparisons are against the wild type or previously dominant variants such as B.1.177.

Table 3: Table showing which combinations of variants we compared.

| Comparison Number | Variant 1 | Variant 2 |
|---|---|---|
| 1 | B.1.177 | B.1.1.7 (Alpha) |
| 2 | B.1.1.7 (Alpha) | B.1.617.2 (Delta) |
| 3 | B.1.617.2 (Delta) | AY.4 (Delta) |
| 4 | B.1.1.7 (Alpha) | B.1.617.2 + AY.4 (Delta) |
| 5 | AY.4 (Delta) | BA.1.1 (Omicron) |
| 6 | B.1.617.2 + AY.4 (Delta) | BA.1.1 + BA.2 (Omicron) |
| 7 | BA.1.1 (Omicron) | BA.2 (Omicron) |

Table 4: Table showing the ΔAIC values for each of the models for the comparisons using raw data. ΔAIC is defined as the AIC value for a specific model, minus the minimum AIC (i.e. that of the best fitting model) across all models. A value of zero is hence given to the best model, with values greater than two usually relating to substantially poorer fit.

| | | ΔAIC | | | | | | |
|---|---|---|---|---|---|---|---|---|
| Comparison Number | | 1 | 2 | 3 | 4 | 5 | 6 | 7 |
| Model | S | 476.8 | 679.7 | 765.7 | 1011.5 | 573.6 | 1078.2 | 557.2 |
| | I | 424.0 | 517.3 | 309.3 | 771.3 | 574.8 | 907.2 | 450.6 |
| | G | 4864.2 | 1559.8 | 18400.3 | 5762.0 | 7114.1 | 9503.3 | 8886.8 |
| | GS | 838.4 | 909.1 | 811.00 | 1024.7 | 536.6 | 979.6 | 846.7 |
| | GI | 0 | 0 | 0 | 0 | 0 | 0 | 0 |
| | Best Fit | GI | GI | GI | GI | GI | GI | GI |

Table 5: Table showing the p-values for the smooth terms in each of the best-fit models fitted to the raw data as reported by the bam summary.

| | | p-value | | | | | | |
|---|---|---|---|---|---|---|---|---|
| Comparison Number | | 1 | 2 | 3 | 4 | 5 | 6 | 7 |
| Smoother | Global | $5.41e^{-05}$ | $3.48e^{-05}$ | $< 2e^{-16}$ | $< 2e^{-16}$ | $< 2e^{-16}$ | $< 2e^{-16}$ | $< 2e^{-16}$ |
| | Variant 1 | $< 2e^{-16}$ | $< 2e^{-16}$ | $< 2e^{-16}$ | $< 2e^{-16}$ | $< 2e^{-16}$ | $< 2e^{-16}$ | $< 2e^{-16}$ |
| | Variant 2 | $< 2e^{-16}$ | $< 2e^{-16}$ | $< 2e^{-16}$ | $< 2e^{-16}$ | $< 2e^{-16}$ | $< 2e^{-16}$ | $< 2e^{-16}$ |

### Results

We present the relative transmission of principal variants normalised by the variant generation time in Figures 3 and 7. These still show a potential for new variants to out-compete the existing dominant variant with a large degree of spatial heterogeneity in realised transmission.

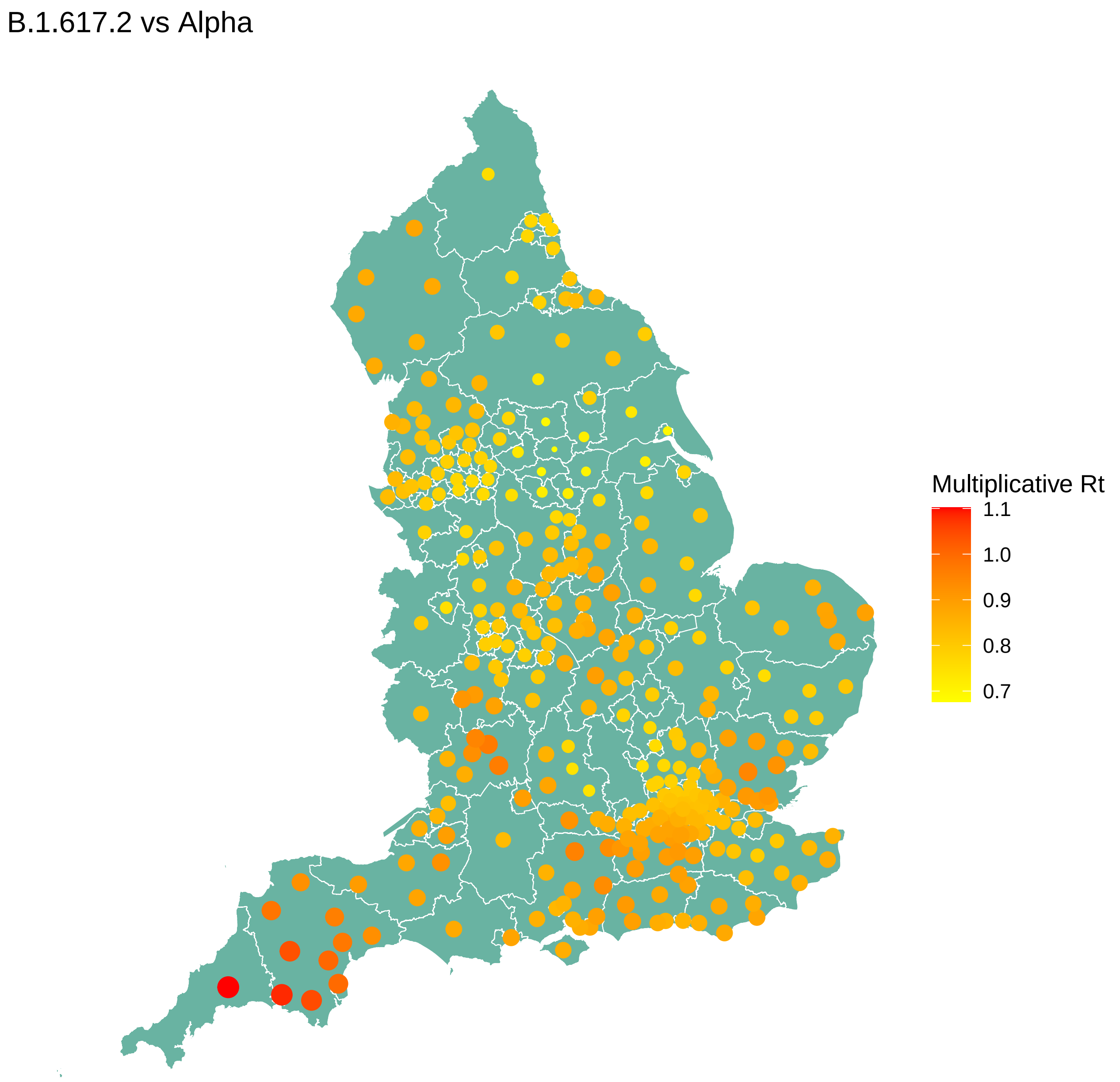


Figure 3: Estimates of relative transmissibility for delta vs alpha, normalised by the difference in generation time of the variant. Generation times used are 5.86 days for Alpha and 5.67 days for Delta [Xu et al., 2023].

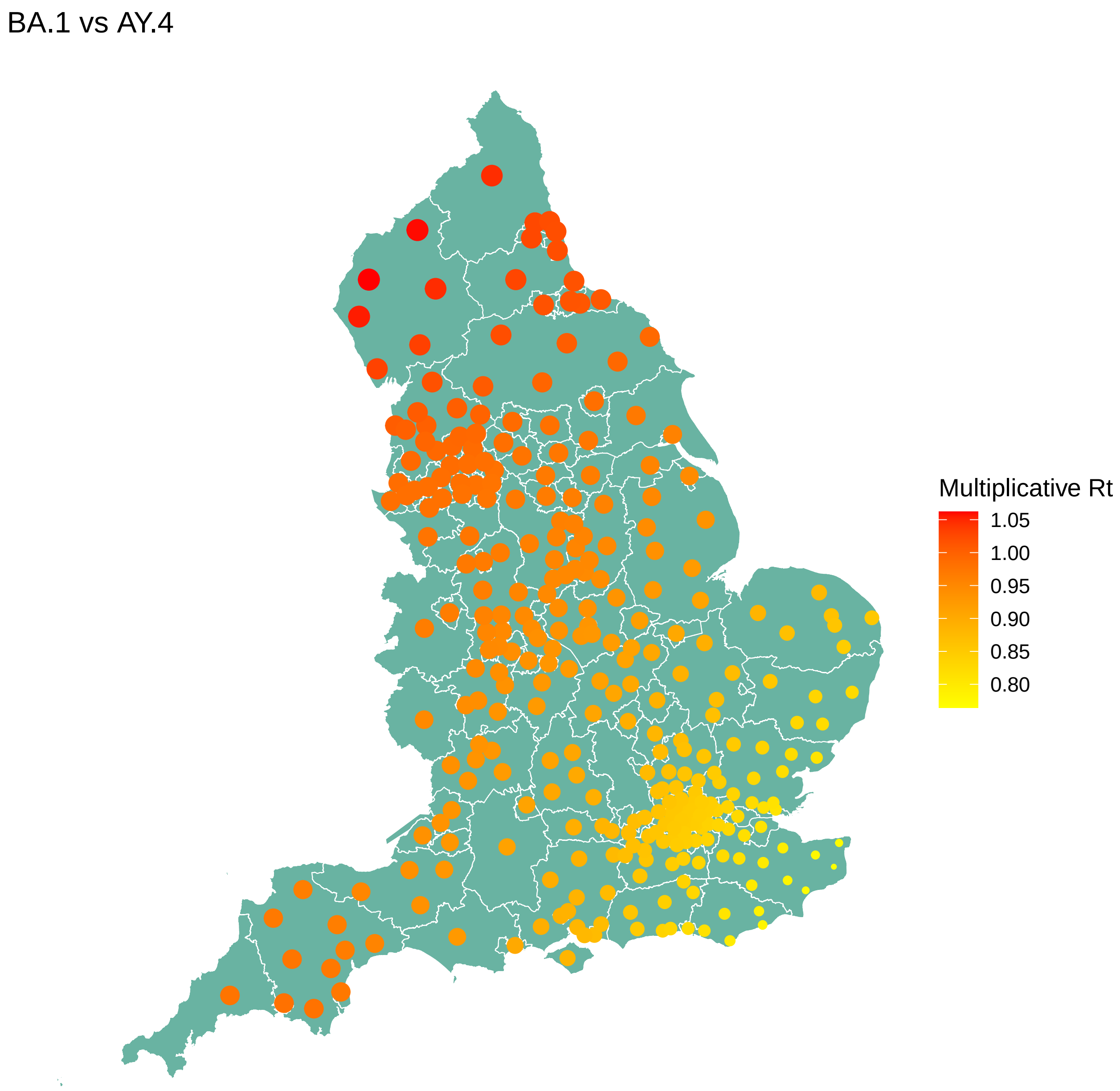


Figure 4: Estimates of relative transmissibility for Omicron BA.1 vs delta AY.4, normalised by the difference in generation time of the variant. Generation times used are 5.67 days for Delta and 6.84 days for Omicron [Xu et al., 2023].

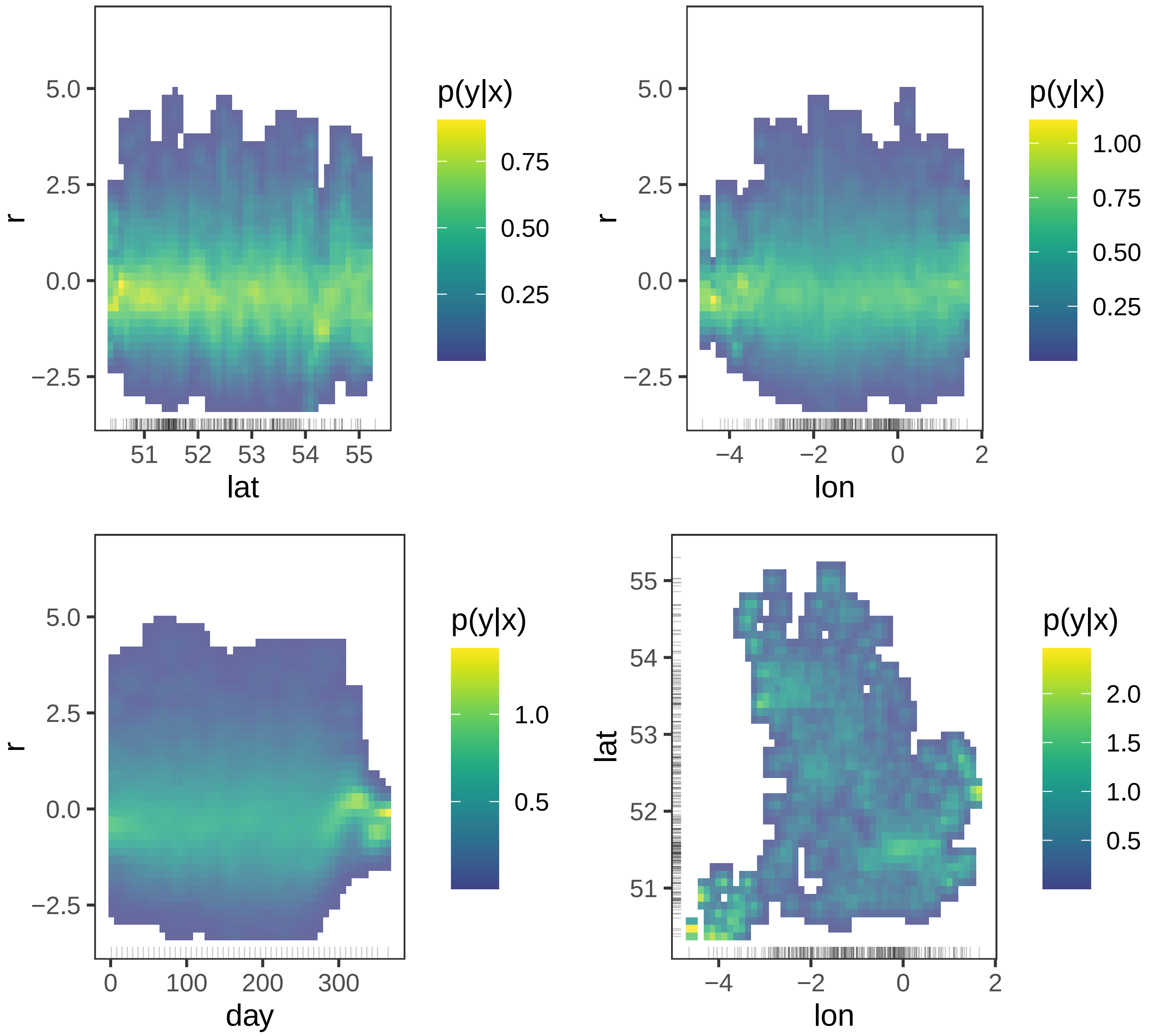


Figure 5: Residual plots across covariates for latitude, longitude and day, as well as a 2D surface of residuals for the alpha vs wild type model. Residuals are largely zero-mean, symmetric and patternless across covariate ranges suggesting a well-fitted model.

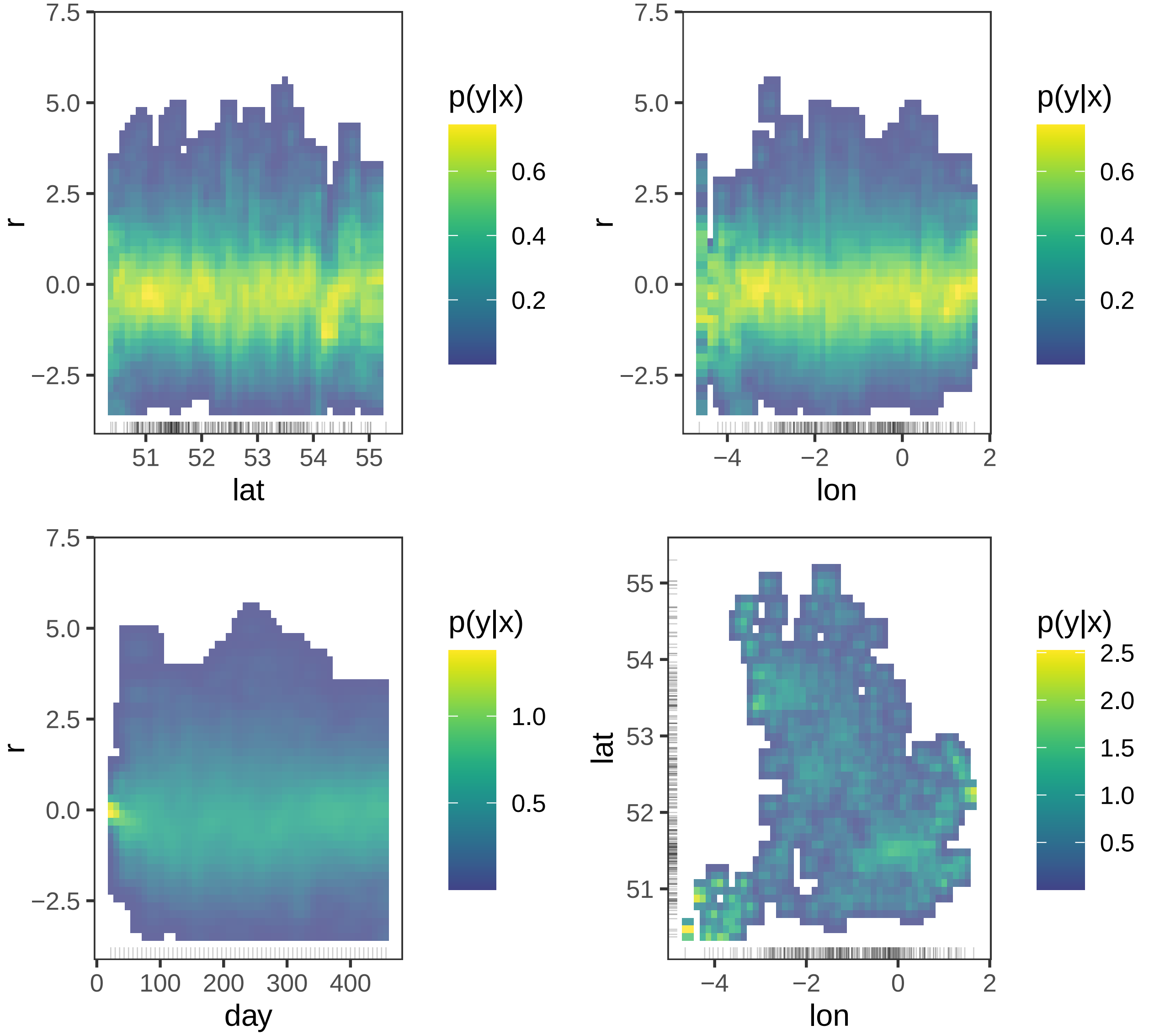


Figure 6: Residual plots across covariates for latitude, longitude and day, as well as a 2D surface of residuals for the delta vs alpha model. Residuals are largely zero-mean, symmetric and patternless across covariate ranges suggesting a well-fitted model.

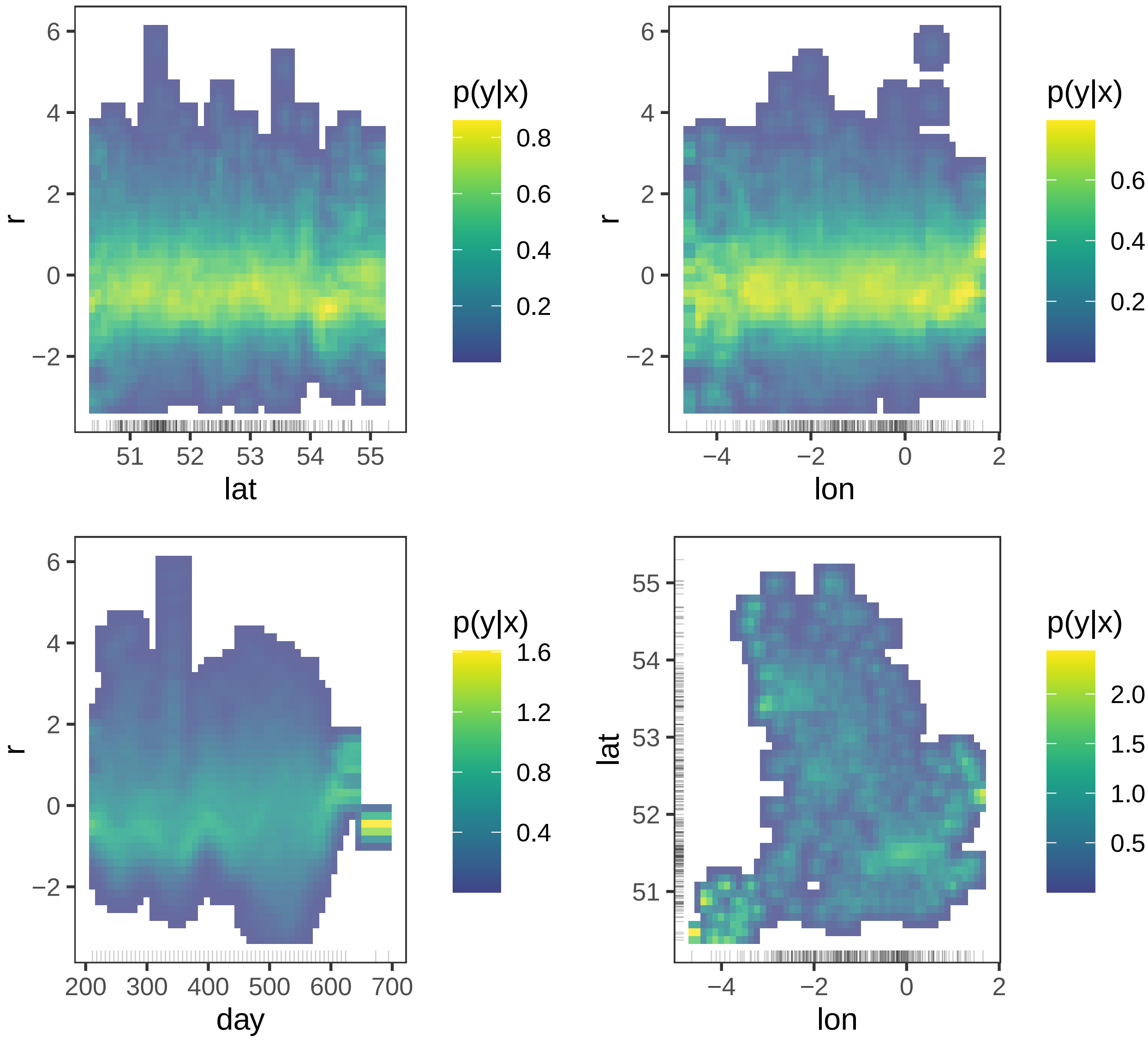


Figure 7: Residual plots across covariates for latitude, longitude and day, as well as a 2D surface of residuals for the delta vs BA.1 model. Residuals are largely zero-mean, symmetric and patternless across covariate ranges suggesting a well-fitted model.

The GAM structure for seropositivity is as follows:

```
sero_mod <- gam(total ~ Lineage+s(day)+s(sero)+
                  t2(sero, day, bs=c("tp", "tp"),by=Lineage, k=c(10, 10), m=2),
                data=sero, method="fREML", family="nb")
```

Table 6: Table of p-values for terms in the GAM.

| GAM term | p-value |
|---|---|
| Lineage | 0.0548 |
| s(day) | 0.8259 |
| s(sero) | 0.0984 |
| t2(sero,day):LineageB.1.1.7 | 0.4494 |
| t2(sero,day):LineageB.1.177 | 0.3436 |
| t2(sero,day):LineageBA.1.1 | 0.6501 |
| t2(sero,day):Lineagedelta | 0.4249 |
